# Why some countries but not others? Urbanisation, GDP and endemic disease predict global SARS-CoV-2 excess mortality patterns

**DOI:** 10.1101/2023.08.06.23293729

**Authors:** Nicholas M. Fountain-Jones, Michael Charleston, Emily Flies, Scott Carver, Luke Yates

## Abstract

The global impact of the SARS-CoV-2 pandemic has been uneven, with some regions experiencing significant excess mortality while others have been relatively unaffected. Yet factors which predict this variation remain enigmatic, particularly at large spatial scales. We used spatially explicit Bayesian models that integrate socio-demographic and endemic disease data at the country level to provide robust global estimates of excess SARS-CoV-2 mortality (P scores) for the years 2020 and 2021. We find that gross domestic product (GDP), spatial patterns and urbanization are strong predictors of excess mortality, with countries characterized by low GDP but high urbanization experiencing the highest levels of excess mortality. Intriguingly, we also observed that the prevalence of malaria and human immunodeficiency virus (HIV) are associated with country-level SARS-CoV-2 excess mortality in Africa and the Western Pacific, whereby countries with low HIV prevalence but high malaria prevalence tend to have lower levels of excess mortality. While these associations are correlative in nature at the macro-scale, they emphasize that patterns of endemic disease and socio-demographic factors are needed to understand the global dynamics of SARS-CoV-2.

## Introduction

Since the emergence of the SARS-CoV-2 pandemic in December 2019, there has been substantial interest in understanding the determinants of disease patterns and accurately assessing the true impact of this worldwide disaster. There have been a wide variety of approaches to estimating the mortality associated with SARS-CoV-2, but in each case, estimates of the true death toll have likely been orders of magnitude higher than reported deaths. Excess mortality, the difference in deaths due to a crisis compared to those expected under normal conditions [1], provides a more reliable estimate of COVID-19 deaths than reported deaths [2]. However, the factors shaping the heterogeneity of estimates of excess deaths across countries remain obscure.

Global estimates for the first epidemic waves in 2020/2021 range from 14.8 [2] to 18.2 million excess deaths [3], dwarfing the 5.94 million reported deaths for the same period. For example, the World Health Organization (WHO) estimated Indonesia to have 1.03 million excess deaths up until December 2021 (credible interval (CI) 0.75-1.29 million) whereas 0.23 million excess deaths were estimated in Pakistan (CI 0.04-0.4 million) [2], even though the population sizes of these countries are comparable. Africa has experienced much lower excess mortality compared to most other regions [2–5]. Deficiencies in the input data in developing countries may underly some of this variation, but these estimates are congruent with cross-sectional studies and reports [e.g., 6].

Comparing excess mortality estimates across countries is a challenging task as raw estimates are strongly dependent on population size and age structure of each country [2,7]. For example, larger populations with higher proportions of older people will have higher number of expected deaths compared to countries with smaller younger populations. To facilitate cross-country comparisons, estimates of excess deaths are often normalized by the expected number of deaths (from all causes) across a period of time. The measurement of excess deaths, referred to as a P-score [7], takes into account both the population size and age structure (see *Methods*). For instance, if 100 deaths were anticipated but the actual number reached 150, resulting in 50 excess deaths, the P-score would be calculated as 50% [7]. Put another way, if a country had a P-score of 50% for a year then the reported death count was 50% higher than the expected death count that year. Recent (as of February 2023) P-score estimates from the WHO have provided important insights into the relative toll of the pandemic across countries, with smaller countries in the Americas suffering relatively more excess deaths (highest P-scores), even though they experienced fewer absolute deaths compared to larger countries [2]. For example, Peru had an estimated P-score of 97% or effectively a doubling of deaths during the first waves of the pandemic [2]. Other countries, such as Australia where infection rates were low were estimated to have negative values as there were actually fewer deaths during these years than expected [2].

Clearly, differences in pandemic response explain some variation in global P-Score estimates but these are not sufficient to explain the general patterns [2]. Here we build a fully probabilistic model accounting for uncertainties in P-score estimates to test if socio-demographic, economic and patterns of endemic disease could help predict these disparate estimates of excess mortality across countries. While P-scores control for age structure of a population, age is a well-known risk factor for mortality, some countries with larger portions of the population > 70 y.o. are likely to experience higher mortality than countries with fewer people in this age bracket [e.g., 8]. As P-scores capture direct and indirect SARS-CoV-2 deaths (e.g., because patients could not access treatment that would be otherwise available), the prevalence of non-communicable conditions such as heart disease [9] may also play a role in shaping these excess deaths. Economic variables such as gross domestic product (GDP) [30] and health spending may also impact mortality rates as countries with high GDP and health spending may lower mortality risks through, for example, better treatment. Countries with high population density and urbanisation may also experience higher mortality as transmission may be facilitated in these conditions [10] or lower, because urban populations tend to have better access to health facilities [11]. Further, our previous work has also found that variation in the prevalence of endemic pathogens including malaria (*Plasmodium* sp.), common intestinal parasitic worms (e.g., *Trichuris trichiura*, the causative agent of ascariasis) and human immunodeficiency virus (HIV) played a surprisingly important role in predicting variation in SARS-CoV-2 reported deaths [12]. For example, countries with relatively high prevalence of endemic malaria had reduced numbers of deaths, whereas countries with higher HIV prevalence had higher numbers of reported deaths (while controlling for other factors such as the mean age of the country) [12].

Spatial relationships are also likely to be important in explaining variation in P-scores and need to be accounted for in global models [e.g., 13]. For example, regions or countries closer in space may have similar P-scores due to similar epidemic trajectories [14]. High connectivity between neighbouring countries could also facilitate the spread of the virus and increase mortality. International air travel is also known to facilitate SARS-CoV-2 spread among countries [13,15,16], and countries with high air travel connectivity with other countries may have increased numbers of introduction events which may result in higher mortality. Countries with high connectivity also tended to have the earliest outbreaks of the virus [17].

In this study, we introduce a Bayesian model-based framework with the aim to discover the potential drivers behind the variations in excess deaths across countries. By utilizing this model-based approach, we illuminate factors that contribute to the diverse severity of the pandemic’s consequences worldwide.

## Methods

### Data retrieval

We extracted P-score estimates from the WHO excess mortality code base on the 3^rd^ of March 2023 (the https://github.com/WHOexcessc19/Codebase). We computed the posterior mean and its standard error for the average P-score estimates across the 24 months spanning 2020 and 2021. We used only data for the first wave of SARS-CoV-2 to examine infection patters without the influence of the variable roll-out of vaccines. We also used the WHO transmission classification scheme (i.e., countries with community transmission, clusters of cases only, sporadic cases and no cases) to account for differences in pandemic response (as of 29^th^ of January 2021, the midpoint of our excess mortality estimates).

We leveraged the predictor variables collated previously [see 12 for further details]. Briefly, we extracted demographic and economic variables from the World Bank dataset (https://data.worldbank.org/) for each country. These variables include per capita GDP (in current USD averaged across 2014-2019), percent of the population living in urban areas (hereafter percent urban). Global health spending estimates (USD) are calculated by the World Health Organization and include healthcare goods and services consumed each year. To analyse the importance of spatial patterns to P-score estimates we constructed a lagged P-score variable based on a neighbourhood matrix. To construct the neighbourhood matrix, we extracted centroid coordinates of countries and generated spatial neighbourhood matrix based on the mean P-score estimate using the *spdep* package in R [18]. To capture the role of country-level connectivity in predicting P-scores, we incorporated the 2007 air connectivity index [16]. We attempted to include an intrinsic spatial conditional autoregressive (iCAR) term to our model, yet we could not attain model convergence with this extra complexity.

We downloaded the mean estimate of prevalence of 14 endemic diseases in each country (Table S1) based on 2019 data from the Institute for Health Metrics and Evaluation (IHME) Global Burden of Disease (GBD) database (https://www.healthdata.org/research-analysis/gbd). As many of the country-level disease prevalence estimates were correlated, we conducted principal component analysis (PCA) and we used the top three resultant orthogonal principal components as predictors in our models. As malaria and HIV were not correlated with any of the principal components, we included these variables separately. To measure overall infectious disease burden, we also extracted estimates of years of life lost (YLLs) for infectious diseases combined in each country in 2019 using the GBD data. We extracted diabetes prevalence and the average cardiovascular death rate for each country in 2019 from the GBD also. See Table S1 for a complete list of predictors used.

We imputed missing data for both datasets dataset using classification and regression trees as implemented in the multivariate imputation by chained equations (MICE) package [19]. We generated ten separate multiple imputations and combined the posterior distributions across the ten corresponding model fits to marginalise over the uncertainty of the replacement values. We screened variables for collinearity by calculating Pearson’s correlation coefficient (ρ) and for correlated pairs excluded the variable with the highest correlation across the complete dataset.

### Bayesian modelling

We modelled P-score variation globally using generalised additive models (GAM) with a Gaussian error distribution to facilitate the inclusion of response errors (i.e., the uncertainty of the P-score estimates) and since the P-scores values are naturally unconstrained. The advantage of GAMs over linear models is their flexibility to capture a wide range of non-linear functional forms using splines while mitigating overfitting through smoothing (we set the maximal basis dimensions of the splines to 6 to flexibly capture non-linear relationships while keeping the effective model complexity under control). Specifically, our models quantified the probability of observing P-scores (*y_i_*) in country *i* given by:

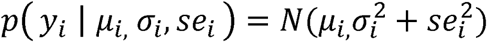

where *se*_i_= *se*(*yi*) is the standard error of the response and µ and cr^2^ are the modelled mean and variance, respectively. Models for each of the distributional parameters are given by:

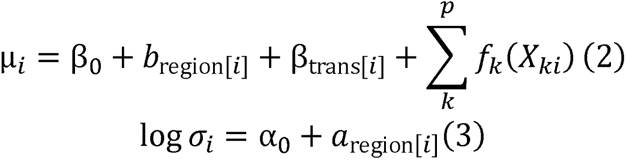

where α_O_and *β*_O_ are global intercepts, a and b are region-level (random) intercepts, β_trans_ is a fixed effect for transmission type, *p* number of predictors included in the model and *f_k_*(*X_kl_*) are smooth spline terms for each continuous predictor *X_k_*. The region-level intercepts (*a*_region[*i*]_, *b*_region[*i*]_) were drawn from a bivariate normal distribution with an estimated three-parameter covariance matrix. The basis elements for each thin plate smoothing spline were generated using the R package *mgcv* [20] and the corresponding regression coefficients were drawn from a normal distribution parametrised by an estimated smoothing hyperparameter.

All models were fit in a Bayesian framework using Hamiltonian Monte Carlo (HMC) methods [21] and ‘no U-turn sampling’ (NUTS, [22]), as implemented in the R package ‘*brms*’ [23]. We ran four chains of 12000 iterations, using weakly informative priors---the prior for the smoothing hyperparameter was half student-t distribution (df = 3, scale = 2.6), and inferences proved robust to changes in the choice of scale. To establish chain convergence, we used the rank-normalised R̂ statistic (R̂< 1.01) [24] as well as visual inspection. We validated the model by comparing pointwise posterior-predictive intervals to the observed P-scores for each country [25].

To interrogate our model, we computed conditional effects for each covariate. For the numeric variables, we computed the estimated change in response (centred on the posterior-predictive mean) as a function of changes in the value of a given predictor (centred on the global mean) while holding all other variables at their global means. Variables were considered strong predictors of P-scores if the 90% credible interval (CI) did not include 0. Variables where this was true for a smaller portion of the range were identified as weaker variables (variables where all values of the CI included 0). To test the utility of adding endemic disease variables to our macroecological models, we evaluated model performance using approximate leave-one-our cross validation (LOO, [26]) an estimate of relative expected Kullback–Leibler discrepancy and compared the model both with and without the inclusion of endemic pathogens. We also compared model performance by region. Our complete workflow and data are available on github: (https://github.com/nfj1380/covid19_macroecology).

## Results

Our model was able to predict global P-score variation remarkably well with mean observed P-scores from 175 out of 181 countries inside the model credible intervals (Fig. 1). Our model underestimated excess mortality for five countries and only overestimated P-scores from Togo. Our model could not predict the P-score for Peru (the highest mean P-score calculated, Fig. 1 inset) but our predictive performance was high for countries with very low or negative P-scores. Our models also underestimated excess mortality in Guam, Oman, Andorra and Armenia (Fig. S1). There was overall no difference in model performance across regions, but countries in the Americas had much higher uncertainty in P-score estimates compared to the other regions. (Fig. 1).

**Fig. 1:**
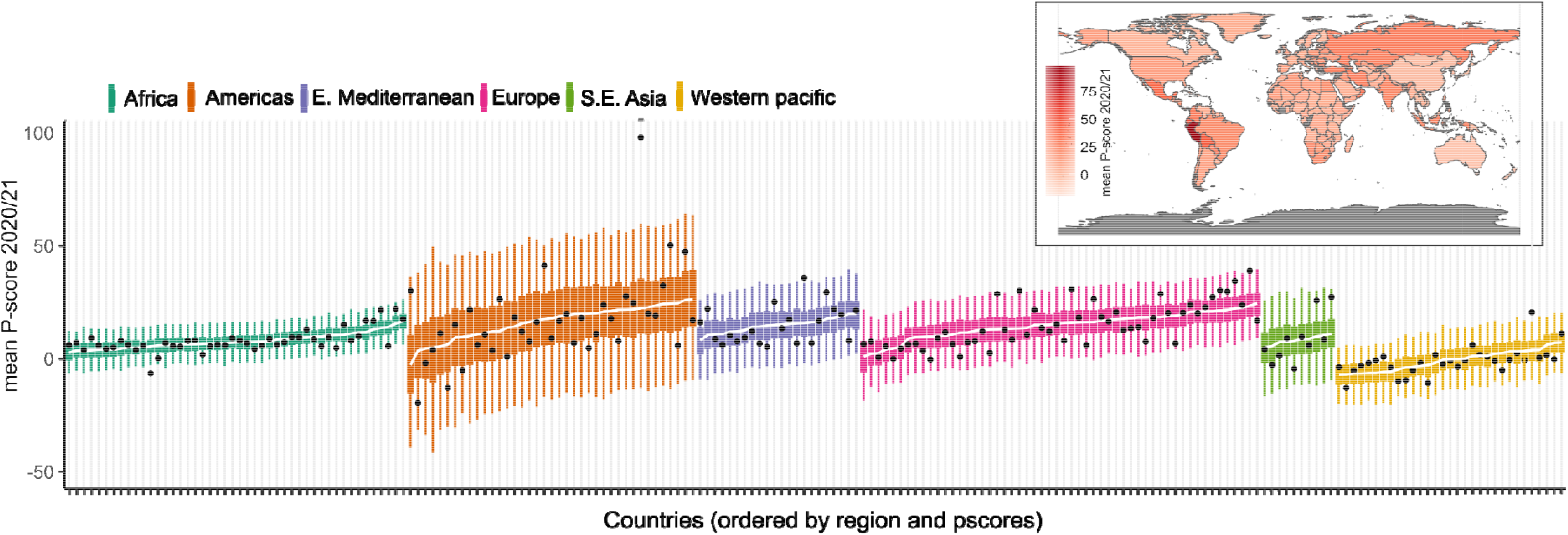
Posterior predictive performance of our P-score model for each country and (inset) the global distribution of mean P-scores acr countries. Countries are grouped by region and ordered by predicted values (lowest to highest). Black dots are the observed values for country and the trend line is the median posterior estimate of the models. Credible intervals (CIs) are coloured by region, with the 50% CIs depicted by thick and thin lines, respectively. See Fig. S1 for predictions with each country labelled.

We found a mix of population, spatial patterns and endemic disease that best predicted P-score variation in our models (Fig. 2). Though important at individual or within country scales, variables such as proportion over 65 y.o., population density and air-connectivity were less important at global scales (Fig. 2a). We also did not see a signature of diabetes prevalence or cardiovascular disease impacting P-score estimates (Fig. 2a, see Fig S2 for the conditional effects plots).

**Fig 2:**
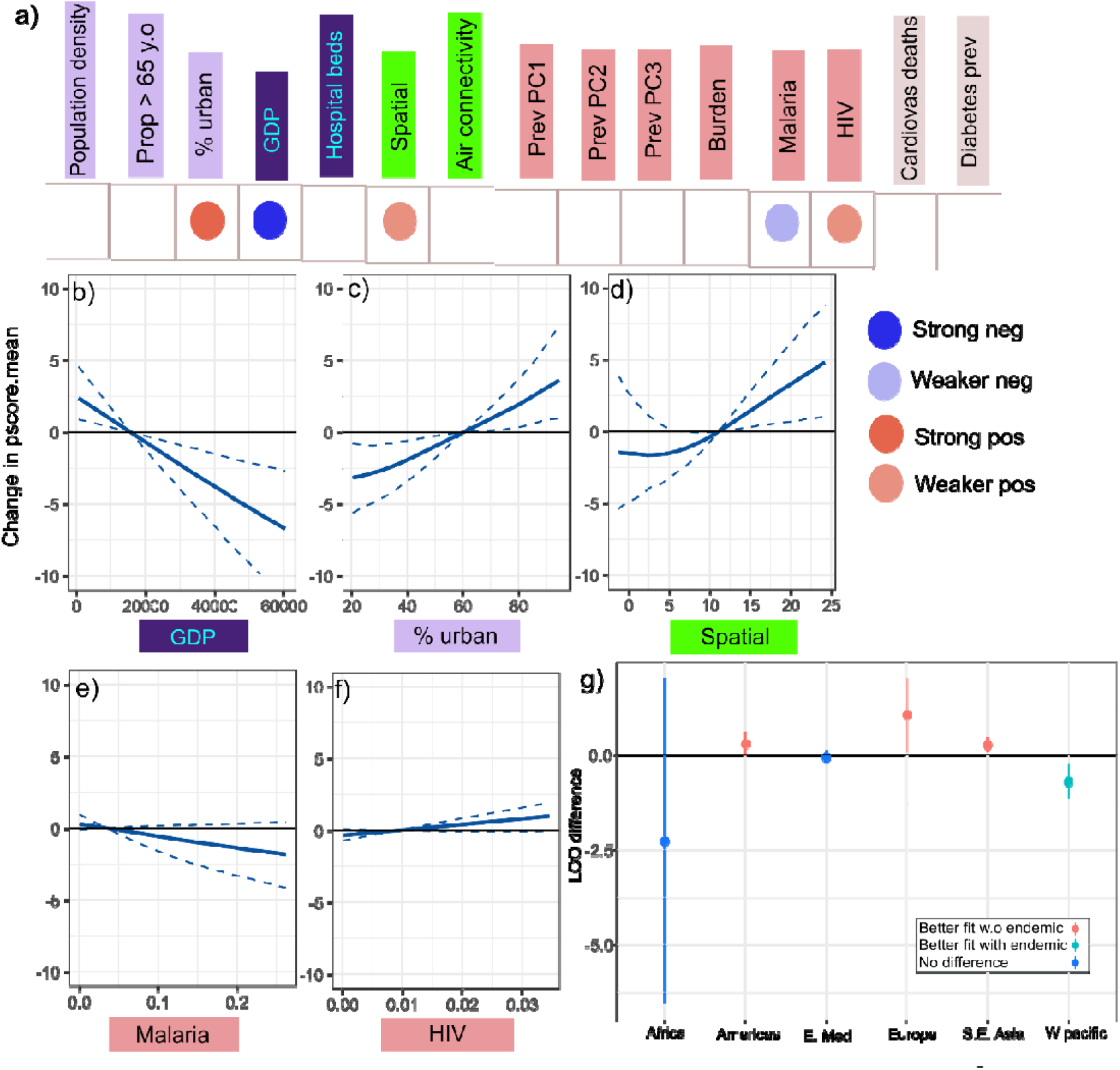
Conditional effects of model predictors on the expected change in global P-scores. a) and direction of covariate effects on P-scores. Pos = positive, Neg = negative. Covariates without a corresponding circle (i.e., blank) had little impact on P-score estimates (see Fig. S2). b-f) Conditional plots showing the expected difference in P-scores as a function of the difference between a given covariate value and the mean observed covariate vales (CIs have a width of 0 at the mean, see *Methods*). span the predictors included in the model. Solid lines denote the posterior means and the associated intervals (dashed lines) denote the 90% credible intervals (CIs). Predictors are colour-coded based on variable type (dark blue = associated with a country’s economic capacity, light red = country level pathogen prevalence estimate, light grey = estimated prevalence of non-infectious disease, light green= spatial variable, purple = population characteristic. Spatial: estimates of cases in neighbouring countries. HIV: Human immunodeficiency virus. (see Table S1 for details). g) Leave one out (LOO) comparisons of our model with all co-variates compared to a model leaving out the endemic disease variables summed across regions. Region level estimates not overlapping the 0 LOO difference were considered significant. W.o = without.

Per-capita GDP and percent urban were strong predictors of P-score variation (Fig. 2a/b). Wealthier countries with a per-capita GDP >$20,000 had decreased P-scores (while controlling for all other predictors in the model, Fig. 2b). GDP decreased excess mortality by up to 10% in countries with a GDP of ∼$60,000 (e.g., USA, Iceland, and Singapore). Conversely, countries with >60% urban population had higher P-scores (Fig. 2b). Spatial patterns played a role and the effect was non-linear (Fig. 2a/b). Countries with neighbours having average P-scores >12 also exhibited higher P-scores (Fig. 2b).

We also detected divergent (but weaker) relationships with malaria and HIV prevalence on P-scores (Fig. 2a). While holding all other variables in the model at their mean value, countries with relatively high prevalence of malaria had decreased P-scores, whereas countries with relatively high HIV prevalence had higher P-scores. The estimated LOO scores indicated that retaining endemic disease variables in the model did not improve model fit overall (ELPD difference = – 1.4, SE = 4.4), although including endemic disease did improve predictions in the Western Pacific (Fig. 2c). There was also some evidence that the fit was improved by endemic disease in Africa but the LOO scores were highly variable which could indicate that we are missing important variables in this region. For Europe and the Americas in contrast, model fit was improved by not including the endemic disease variables (i.e., LOO model difference scores above 0). Collectively, there appears some regional scale context-dependent relationships between SARS-CoV-2 P-scores and endemic disease.

## Discussion

Our study provides some important insights into the divergent impacts of the SARS-CoV-2 pandemic. Our Bayesian analytical approach combined with robust estimates of excess deaths from the WHO revealed that GPD, urbanisation and spatial patterns played an important role in predicting excess mortality across countries. We also found that malaria and HIV were associated with excess deaths, particularly in the Western Pacific and African regions, but not necessarily in other regions of the world. Studies at this scale, while necessarily associative, can help identity plausible drivers of variation in SARS-CoV-2 mortality at large spatial scales.

We identified that highly urban countries with low GDP and surrounded by countries with high P-Scores had the highest levels of excess mortality during the first waves of the pandemic. Endemic disease patterns also had some association, but comorbidities such as diabetes prevalence and proportion of the population > 65 y.o. were less relevant at the scale of our analysis. The relationship between GDP and SARS-CoV-2 mortality has generally been found to be positively correlated with the number of reported deaths higher in countries with higher GDP [e.g., 27]. Our model using the more robust P-score estimates of excess mortality suggest the opposite trend; potentially as our estimates are less-impacted by reporting biases in countries with fewer resources. GDP is strongly correlated with health spending, so it is possible that the quality of health care afforded by countries with high GDP is associated with reduced excess mortality. However, even after accounting for GPD, the burden of excess mortality is not shared evenly. Highly urban populations are known to be vulnerable to SARS-CoV-2 mortality as urban areas are densely populated and are travel hubs, both of which facilitate transmission [10,12,16,27]. Despite the benefits of living in cities, such as better access to medical resources, our study demonstrates that urban areas face amplified SARS-CoV-2-related mortality [11].

Highly urban countries are often more connected to other countries, yet higher air connectivity alone did not explain P-score variation in our model. Spatial relationships as well as regional variability were more important and similar results have been found in models focusing on global patterns of confirmed deaths [12]. In our analysis, a clear threshold emerged indicating a spatial effect, where mean P-scores increased when the average of neighbouring countries exceeded approximately 10. This finding suggests geographically constrained patterns, with countries experiencing high transmission and mortality influencing neighbouring P-scores through cross-border transmission.

We also provide further evidence for the effect of endemic pathogen prevalence on global SARS-CoV-2 mortality with countries with lower malaria prevalence but higher HIV prevalence tending to have higher P-scores. While the effect at a global level was marginal, our interrogation of regional level patterns found that endemic pathogens were associated with P-score patterns in the regions with high burdens of endemic disease. Our findings are similar to other studies modelling deaths at this scale and our previous work [12,16]. In these regions, coinfections with either of these pathogens and SARS-CoV-2 were common and coinfection can impact the severity of SARS-CoV-2 [28–30]. For example, immunocompromised HIV patients can face a higher risk of severe SARS-CoV-2 infection [31]. At a coarser scale, countries in Africa with high-levels of mortality also had the highest prevalence of HIV [16]. Additionally, there is evidence suggesting that malaria can influence the mortality rates of SARS-CoV-2 by mitigating the severity of the illness [29,32]. For instance, a study conducted on healthcare workers in India revealed that individuals coinfected with malaria and SARS-CoV-2 experienced an average recovery period that was eight days shorter than those who were infected with SARS-CoV-2 alone [29].

Though our model for the most part could predict country-level P-scores well, it is important to acknowledge that this was not the case for all countries. For example, the P-score for Peru exceeded our model estimates. Why Peru is globally unique is unclear, but may be linked to failures in the health system coupled with diverse populations with high levels of poverty [33]. Although we gathered a diverse range of predictors, the dataset is not comprehensive, and we may have missed important axes of variation. Our objective was to include as many countries as possible in the analysis while minimizing missing data. Our data was also aggregated at the country-level and we possibly missed important variation within countries that may provide high resolution predictions of SARS CoV-2 mortality. Moreover, while our model was able to propagate the estimated uncertainty with the WHOs modelled P-scores, it is important to recognise some features of WHOs generation of the P-scores themselves. The WHO found that tracking excess mortality directly in some parts of the world, particularly in sub-Saharan Africa, was not possible and within-country estimates relied on statistical models [2]. We urge caution when interpreting our cross-country models in the region. Still as reported deaths data are likely a severe undercount in sub-Saharan Africa [2,34], model-based P-score estimates provide the most reliable estimates of SARS-CoV-2 mortality in the region.

Our study highlights factors associated with variation in excess mortality across countries and provides insights into why some countries were impacted more by the pandemic than others. By understanding the predictors of P-score variation across countries and gaining insights into the differential impacts of the pandemic, we may be able to better inform global strategies for outbreak management and response. For example, our model estimates suggest that targeting medical resources to highly urban countries with low GDP and high HIV prevalence may reduce mortality during future outbreaks. Future investigations should aim to explore these global factors using increasingly accurate mortality estimates and consider the dynamics of new waves of SARS-CoV-2.

## Data availability statement

All data and code used to perform the analyses presented in this paper are available on Github: https://github.com/nfj1380/covid19_macroecology

## Acknowledgements

This project was supported by an Australian Research Council Discovery Project Grant (DP190102020).

## Conflicts of interest

None to declare

## Notes

### Competing Interest Statement

The authors have declared no competing interest.

## References

1. Checchi F, Roberts L. In press. A primer for non-epidemiologists.

2. Msemburi W, Karlinsky A, Knutson V, Aleshin-Guendel S, Chatterji S, Wakefield J. 2023 The WHO estimates of excess mortality associated with the COVID-19 pandemic. Nature 613, 130–137. (doi:10.1038/s41586-022-05522-2)

3. Wang H et al. 2022 Estimating excess mortality due to the COVID-19 pandemic: a systematic analysis of COVID-19-related mortality, 2020–21. The Lancet 399, 1513–1536. (doi:10.1016/S0140-6736(21)02796-3)

4. 2023 The pandemic’s true death toll. The Economist. See https://www.economist.com/graphic-detail/coronavirus-excess-deaths-estimates (accessed on 24 May 2023).

5. Bouba Y, Tsinda EK, Fonkou MDM, Mmbando GS, Bragazzi NL, Kong JD. 2021 The Determinants of the Low COVID-19 Transmission and Mortality Rates in Africa: A Cross-Country Analysis. Front Public Health 9, 751197. (doi:10.3389/fpubh.2021.751197)

6. Salyer SJ et al. 2021 The first and second waves of the COVID-19 pandemic in Africa: a cross-sectional study. The Lancet 397, 1265–1275. (doi:10.1016/S0140-6736(21)00632-2)

7. In press. A pandemic primer on excess mortality statistics and their comparability across countries. Our World in Data. See https://ourworldindata.org/covid-excess-mortality (accessed on 24 May 2023).

8. Sorci G, Faivre B, Morand S. 2020 Explaining among-country variation in COVID-19 case fatality rate. Sci Rep 10, 18909. (doi:10.1038/s41598-020-75848-2)

9. Han L, Zhao S, Li S, Gu S, Deng X, Yang L, Ran J. 2023 Excess cardiovascular mortality across multiple COVID-19 waves in the United States from March 2020 to March 2022. Nat Cardiovasc Res 2, 322–333. (doi:10.1038/s44161-023-00220-2)

10. González-Val R, Sanz-Gracia F. 2022 Urbanization and COVID-19 incidence: A cross-country investigation. Papers in Regional Science 101, 399–415. (doi:10.1111/pirs.12647)

11. Scheil-Adlung X. 2015 Global evidence on inequities in rural health protectionLJ: new data on rural deficits in health coverage for 174 countries. ILO Working Papers

12. Fountain-Jones NM, Yates L, Flies E, Flies A, Carver S, Charleston M. 2021 Patterns of SARS-CoV-2 exposure and mortality suggest endemic infections, in addition to space and population factors, shape dynamics across countries. medRxiv, 2021.07.12.21260394. (doi:10.1101/2021.07.12.21260394)

13. Krisztin T, Piribauer P, Wögerer M. 2020 The spatial econometrics of the coronavirus pandemic. Lett Spat Resour Sci 13, 209–218. (doi:10.1007/s12076-020-00254-1)

14. Lacasa L, Challen R, Brooks-Pollock E, Danon L. 2020 A flexible method for optimising sharing of healthcare resources and demand in the context of the COVID-19 pandemic. PLOS ONE 15, e0241027. (doi:10.1371/journal.pone.0241027)

15. Zhong L, Diagne M, Wang W, Gao J. 2021 Country distancing increase reveals the effectiveness of travel restrictions in stopping COVID-19 transmission. Commun Phys 4, 1–12. (doi:10.1038/s42005-021-00620-5)

16. Zhang F et al. 2021 Predictors of COVID-19 epidemics in countries of the World Health Organization African Region. Nat Med 27, 2041–2047. (doi:10.1038/s41591-021-01491-7)

17. Bickley SJ, Chan HF, Skali A, Stadelmann D, Torgler B. 2021 How does globalization affect COVID-19 responses? Globalization and Health 17, 57. (doi:10.1186/s12992-021-00677-5)

18. Bivand R, Piras G. 2015 Comparing Implementations of Estimation Methods for Spatial Econometrics. Journal of Statistical Software 63, 1–36. (doi:10.18637/jss.v063.i18)

19. Azur MJ, Stuart EA, Frangakis C, Leaf PJ. 2011 Multiple imputation by chained equations: what is it and how does it work? Int J Methods Psychiatr Res 20, 40–49. (doi:10.1002/mpr.329)

20. Wood S. In press. Generalized Additive Models: An Introduction with R, Second Edition -. See https://www.routledge.com/Generalized-Additive-Models-An-Introduction-with-R-Second-Edition/Wood/p/book/9781498728331 (accessed on 8 July 2021).

21. Neal RM. 2011 MCMC using Hamiltonian dynamics. arXiv:1206.1901 *[physics, stat]* (doi:10.1201/b10905)

22. Homan MD, Gelman A. 2014 The No-U-turn sampler: adaptively setting path lengths in Hamiltonian Monte Carlo. J. Mach. Learn. Res. 15, 1593–1623.

23. Bürkner P-C. 2017 brms: An R Package for Bayesian Multilevel Models Using Stan. Journal of Statistical Software 80, 1–28. (doi:10.18637/jss.v080.i01)

24. Vehtari A, Gelman A, Simpson D, Carpenter B, Bürkner P-C. 2021 Rank-Normalization, Folding, and Localization: An Improved R̂ for Assessing Convergence of MCMC. Bayesian Analysis 1, 1–38. (doi:10.1214/20-BA1221)

25. Gabry J, Simpson D, Vehtari A, Betancourt M, Gelman A. 2019 Visualization in Bayesian workflow. Journal of the Royal Statistical Society: Series A (Statistics in Society) 182, 389–402. (doi:10.1111/rssa.12378)

26. Vehtari A, Gelman A, Gabry J. 2017 Practical Bayesian model evaluation using leave-one-out cross-validation and WAIC. Stat Comput 27, 1413–1432. (doi:10.1007/s11222-016-9696-4)

27. Skórka P, Grzywacz B, Moroń D, Lenda M. 2020 The macroecology of the COVID-19 pandemic in the Anthropocene. PLOS ONE 15, e0236856. (doi:10.1371/journal.pone.0236856)

28. Bradbury RS, Piedrafita D, Greenhill A, Mahanty S. 2020 Will helminth co-infection modulate COVID-19 severity in endemic regions? Nat Rev Immunol 20, 342. (doi:10.1038/s41577-020-0330-5)

29. Mahajan NN et al. 2021 Co-infection of malaria and early clearance of SARS-CoV-2 in healthcare workers. Journal of Medical Virology 93, 2431–2438. (doi:https://doi.org/10.1002/jmv.26760)

30. Miguel DC, Brioschi MBC, Rosa LB, Minori K, Grazzia N. 2021 The impact of COVID-19 on neglected parasitic diseases: what to expect? Trends in Parasitology 37, 694–697. (doi:10.1016/j.pt.2021.05.003)

31. Huang J et al. 2020 Epidemiological, virological and serological features of COVID-19 cases in people living with HIV in Wuhan City: A population-based cohort study. *Clin Infect Dis*, ciaa1186. (doi:10.1093/cid/ciaa1186)

32. Achan J et al. 2022 Current malaria infection, previous malaria exposure, and clinical profiles and outcomes of COVID-19 in a setting of high malaria transmission: an exploratory cohort study in Uganda. The Lancet Microbe 3, e62–e71. (doi:10.1016/S2666-5247(21)00240-8)

33. Taylor L. 2021 Covid-19: Why Peru suffers from one of the highest excess death rates in the world. BMJ 372, n611. (doi:10.1136/bmj.n611)

34. Cabore JW et al. 2022 COVID-19 in the 47 countries of the WHO African region: a modelling analysis of past trends and future patterns. The Lancet Global Health 10, e1099–e1114. (doi:10.1016/S2214-109X(22)00233-9)

