## Supplementary materials for "Why some countries but not others? Urbanisation, GDP and endemic disease predict global SARS-CoV-2 excess mortality patterns"

**--Supplementary information**

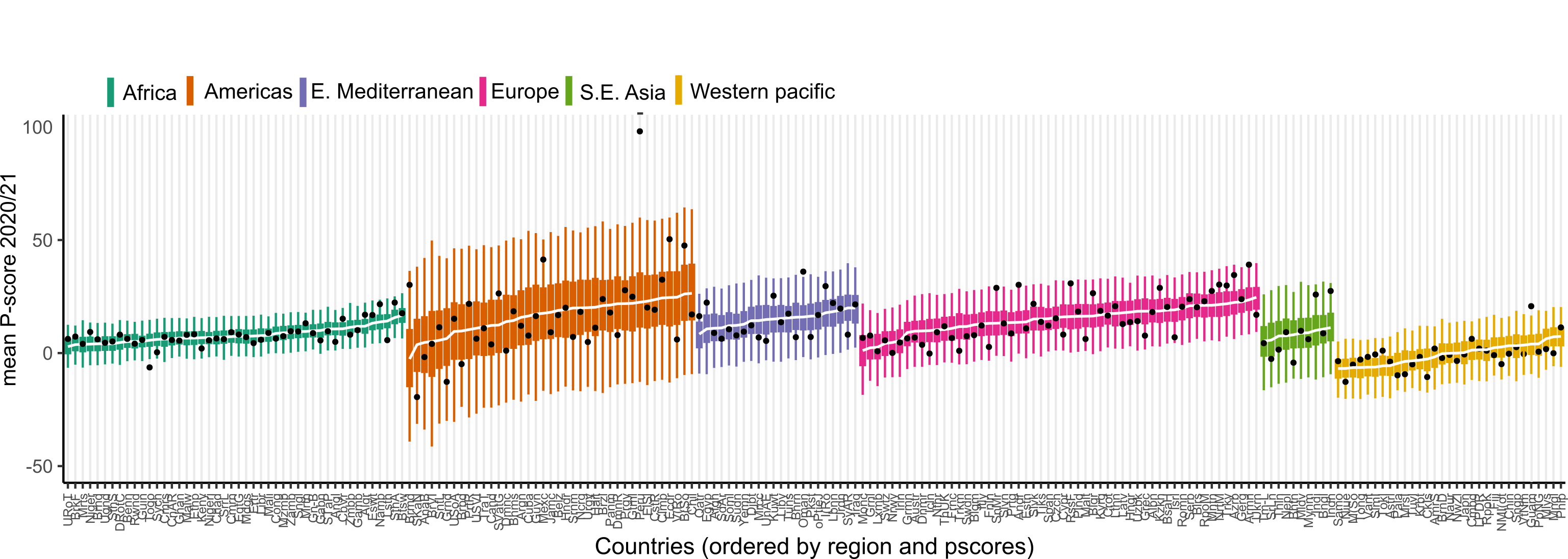

**Fig. S1:** Predictive performance of our P-score model--- for each country. Countries are ordered by predicted values (lowest to highest). CIs are coloured by region. Afri: Africa, Amer = Americas, EMed = Eastern Mediterranean, SEAs = South East-Asia, WPac = Western Pacific. See Appendix S1 for country acronym

**Table S1:** Summary of predictors used to construct our models. See Fig S3 for the correlation plot.

| **Predictor** | **Type** | **Data Source (global)** | **Units** | **Notes** |
| --- | --- | --- | --- | --- |
| Population density | Pop | Average from 2015-19: <https://data.worldbank.org/> | People per km^2^ |  |
| Proportion  > 65 y.o | Pop | Average from 2015-19: <https://data.worldbank.org/> | % |  |
| % urban | Pop | Average from 2015-19: <https://data.worldbank.org/> | % |  |
| GDP | E | Average from 2015-19: <https://data.worldbank.org/> | Per-capita GPD |  |
| Health spending | E | Average from 2015-19: <https://data.worldbank.org/> | US dollars |  |
| Hospital beds | E | Average from 2015-19: <https://data.worldbank.org/> | Beds per-capta | Strongly correlated to health spending* |
| Herpes* | P | Institute for Health Metrics and Evaluation (IHME) GBD database: <https://vizhub.healthdata.org/gbd-compare/> | Estimated prevalence |  |
| Human immunodeficiency virus (HIV) *^#^ | P | Institute for Health Metrics and Evaluation (IHME) GBD database: <https://vizhub.healthdata.org/gbd-compare/> | Estimated prevalence |  |
| Malaria*^#^ | P | Institute for Health Metrics and Evaluation (IHME) GBD database: <https://vizhub.healthdata.org/gbd-compare/> | Estimated prevalence |  |
| Tuberculosis (TB)* | P | Institute for Health Metrics and Evaluation (IHME) GBD database: <https://vizhub.healthdata.org/gbd-compare/> | Estimated prevalence |  |
| Ascariasis* | P | Institute for Health Metrics and Evaluation (IHME) GBD database: <https://vizhub.healthdata.org/gbd-compare/> | Estimated prevalence |  |
| Hookworm* | P | Institute for Health Metrics and Evaluation (IHME) GBD database: <https://vizhub.healthdata.org/gbd-compare/> | Estimated prevalence |  |
| Schistosomiasis* | P | Institute for Health Metrics and Evaluation (IHME) GBD database: <https://vizhub.healthdata.org/gbd-compare/> | Estimated prevalence |  |
| Trichuriasis* | P | Institute for Health Metrics and Evaluation (IHME) GBD database: <https://vizhub.healthdata.org/gbd-compare/> | Estimated prevalence |  |
| Lymphatic filariasis* | P | Institute for Health Metrics and Evaluation (IHME) GBD database: <https://vizhub.healthdata.org/gbd-compare/> | Estimated prevalence |  |
| Infectious disease burden | P | Institute for Health Metrics and Evaluation (IHME) GBD database: <https://vizhub.healthdata.org/gbd-compare/> | Years of life lost (YLLs) |  |
| Diabetes | C | Our world in data (2017 estimate): <https://ourworldindata.org/> | Prevalence |  |
| Cardiovascular deaths | C | Our world in data (2017 estimate): <https://ourworldindata.org/> | Prevalence |  |
| Spatial network | Sp | Network created using the centroid coordinates of each country (see *Methods*) | Cases in countries close in space |  |
| Air travel | Sp | Arvis, et al., .2011 | Air connectivity index (2011) | Strongly correlated with GDP*. |

*Variables collinear and dimension reduction employed using principal component analysis (PCA). ^#^: Variable not correlated with PCA eigenvectors so added separately. S: Direct predictor of SARS-CoV-2 dynamics. Pop: Population characteristic, E: Economic variable, Cl: climate data, P: Measure of human endemic disease, C: Chronic disease prevalence, Sp: Spatial component. ^: Used only in the death model. * See Fig. S2 for correlations.

**Reference**

Hasell J, Mathieu E, Beltekian D, Macdonald B, Giattino C, Ortiz-Ospina E, Roser M, Ritchie H. 2020 A cross-country database of COVID-19 testing. Scientific Data 7, 345. (doi:10.1038/s41597-020-00688-8)

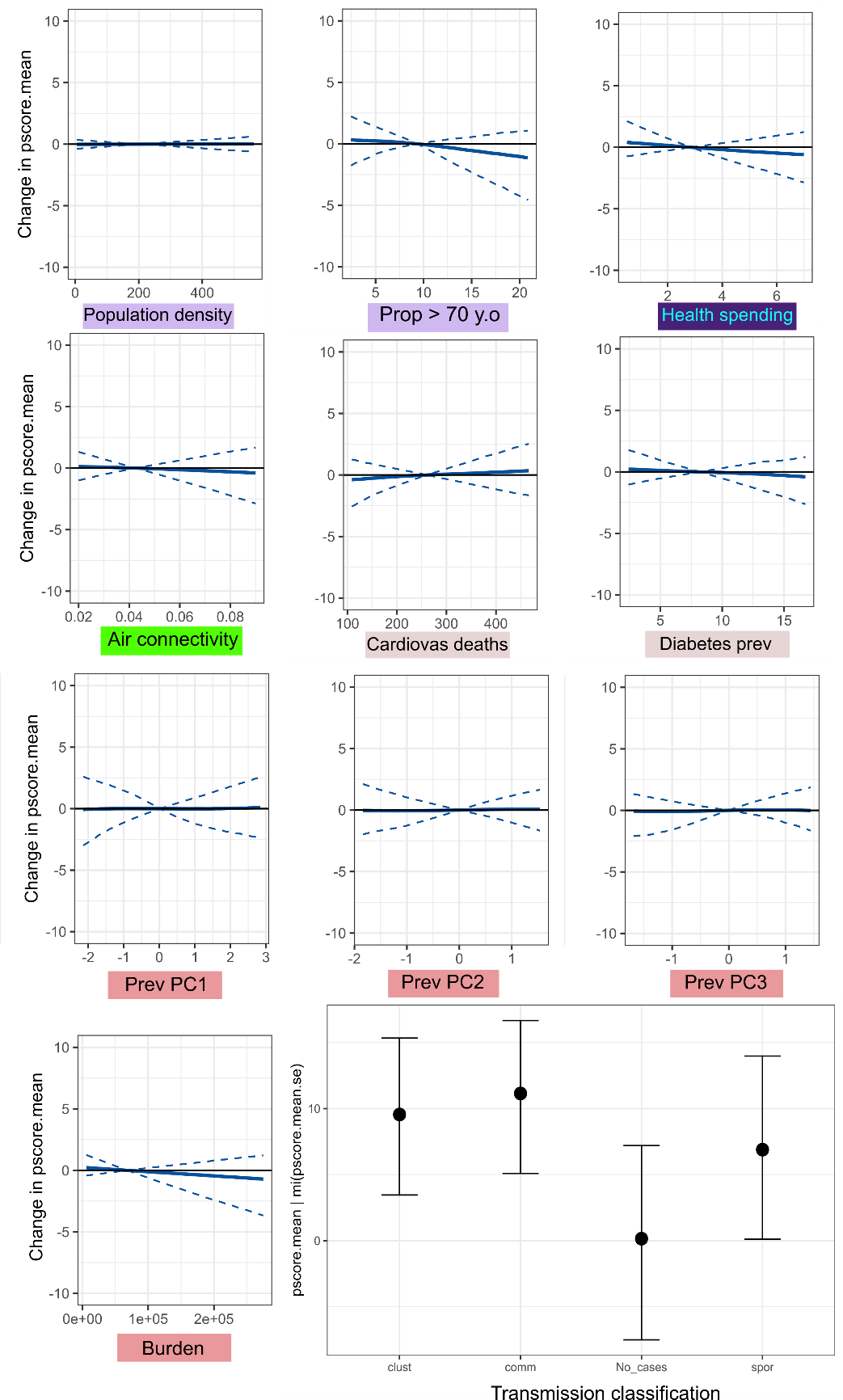

**Fig S2:** Conditional effect plots for other variables with little effect of P-scores in the model (see Figure 2 in the main text for the remaining variables). Predictors are colour-coded based on variable type (dark blue = associated with a country’s economic capacity, light red = country level pathogen prevalence estimate, light grey = estimated prevalence of non-infectious disease, light green= spatial variable, purple = population characteristic. Clust: clustered, comm: community transmission, spor: sporadic cases.

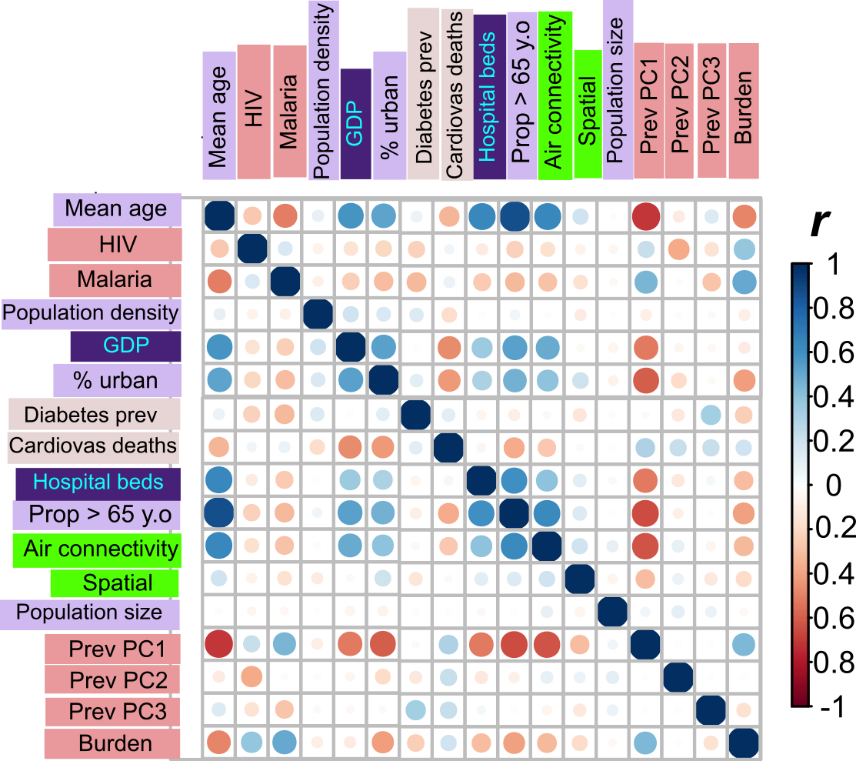

**Fig. S3**: Pearson correlation heat map of the correlations between predictors in the global model. Variables where ρ > 0.7 and had higher overall mean ρ values across all other variables were removed from the analysis.
